# Statistical models of hospital patient fatality rates after accidental falls in children

**DOI:** 10.1101/2023.12.12.23299626

**Authors:** Cyrille Rossant, Leila Schneps

## Abstract

Pediatric traumatic brain injuries represent a public health concern globally. Epidemiological studies are essential to understand their extent, uncover risk factors and outcome predictors, and establish efficient prevention policies. Etiological investigations also benefit from epidemiological data. In young, pre-linguistic children, distinguishing between accidental and non-accidental trauma is often challenging for frontline clinicians. Classification errors can have severe repercussions for children and their families. Past influential epidemiological studies have drawn general conclusions about the etiology of pediatric traumatic brain injuries based on specific statistical information, such as hospital patient fatality rates after reported accidental falls. In this article, we use simple mathematical models to revisit one of these studies and discuss the reliability of its conclusion. We show that apparent paradoxical discrepancies, such as hospital patient fatality rates that are higher after short versus long falls, may have mathematical justification and do not necessarily justify seeking alternative etiological explanations such as non-accidental trauma.

## 1. Introduction

Traumatic brain injuries (TBI) occur when significant external forces are applied to the skull and brain, resulting in potentially severe neurological injuries or even death. In the United States (US) alone, more than 600,000 consultations to emergency departments concern children suffering from TBI [1], making it the leading cause of child death [2]. Globally, three million children are affected annually [3].

Since the adoption of mandatory reporting statutes in the US [4], healthcare professionals have developed protocols to detect suspicions of child abuse, such as non-accidental trauma. Distinguishing between pediatric injuries caused by accidents or abuse is essential to protect a child from an unsafe environment or prevent unjustified legal procedures with long-lasting consequences for the child and its family. However, it is medically challenging for several reasons. First, there is an infinite variety of possible injury mechanisms in both accidental and non-accidental situations. Second, the laws of physics do not depend on the intention behind a particular traumatic event, and inferring intent from medical observations alone is virtually impossible. Third, pre-linguistic children are unable to indicate the circumstances of a traumatic event, and healthcare providers must rely on the interrogation of parents or caregivers.

In cases of suspected non-accidental trauma, the reported clinical history may not always be truthful for obvious reasons. Discrepancies between a reported trivial accidental trauma and severe injuries in a child have historically been considered as a marker of nonaccidental etiologies [5].

Of particular importance is the question of severe or fatal brain injuries after short accidental falls in children. In this age group, household falls are extremely common, representing approximately half of all infants [6]. Premobile infants may be accidentally dropped or fall from the changing table, the sofa, or the bed. As they start crawling, sitting, and standing up, they may fall from their height, with their head potentially impacting a hard surface.

The likelihood of severe injuries depends on a wide range of factors: the type of surface, the age, height, and weight of the child, the kinematics of the fall (the potential for severe injuries after head-first and feet-first falls is clearly not similar [7]), potentially existing inherent risk factors and fragilities of the child, among other factors.

Can such generally benign household falls cause severe or fatal injuries in children? If so, how often? When a story of a short fall is provided at the hospital by parents or caretakers to explain severe injuries of a child, how can one determine whether the story is truthful? Under what conditions should non-accidental trauma be suspected?

To address these challenging questions, epidemiological studies have been conducted. Relying on population statistics may allow one to draw general conclusions about the severity of injuries after household accidental trauma. A particularly influential and frequently-cited (both in medical articles and in medical expert reports) study revealed a major discrepancy in the hospital patient fatality rate after alleged accidental falls between short falls and long falls [8]. In this study, the authors included all children admitted to the Trauma Center at Children’s Hospital San Diego between August 1984 and March 1988 with a history of a “fall”. Among 317 cases, there were 100 short falls (30–120 cm) and 118 long falls (3-13 m). There were 7 deaths in the first group and only 1 death in the latter group. This surprising result was interpreted as follows: *“if the histories of short falls are accepted as correct, the conclusion would be reached that the risk of death is eight times greater in children who fall from 1 to 4 feet than for those who fall from 10 to 45 feet. Since this conclusion appears absurd, it is necessary to seek another explanation for the observed relationship*.*”* To the authors, *“the best explanation of the finding is that for the seven children who died following short falls the history was falsified*.*”* Their general conclusion is that *“when children incur fatal injuries in falls of less than 4 feet, the history is incorrect*.*”*

This study was examined in [9, 10]. In this article, we use a simple probabilistic model to revisit this apparent statistical paradox and investigate key confounding variables that might naturally explain this discrepancy without necessarily assuming that all histories of short falls are falsified.

### 2. Methods

We present a simple probabilistic model that represents the occurrence of accidental falls in children, potentially resulting in fatal injuries and leading, in some cases, to hospital consultations. The objective of the model is to investigate the hospital patient fatality rate of children after falls of various heights and examine whether this hospital patient fatality rate is an increasing function of the fall height.

Following [8], we use the term *short falls* to refer to accidental falls of children from a maximal height of 1.20 m, and *long falls* to refer to falls from a distance greater than 3 meters. For simplicity, we do not consider falls from an intermediate height.

Within a given time period, a child may experience a short fall with probability *f*_*s*_ or a long fall with probability *f*_*l*_. The child may eventually develop fatal injuries with probability *d*_*s*_ after a short fall and *d*_*l*_ after a long fall. Furthermore, we assume, for simplicity, that nonfatal cases do not involve any severe symptoms.

We assume that, after a fall, all children will be brought to the hospital, *except* after short, nonfatal falls. Critically, in these cases, children are brought to the hospital with probability *h*. This assumption takes into account the fact that benign short falls in children are highly common but rarely lead to a consultation at the hospital. On the other hand, long falls are likely to be much rarer, usually involving falls from playground equipments or apartment windows. Even if a child may not appear to be severely injured, a consultation at the hospital is practically certain (in the model, we assume that it is always the case).

We assume all deaths occur at the hospital (which may not be the case in reality, as mentioned in the discussion). Also, all variables are conditionally independent. Figure 1 shows a graphical probabilistic representation of this model.

**Figure 1:**
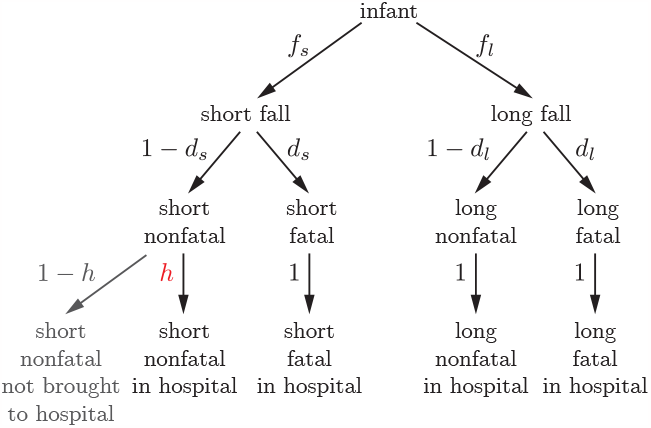
Graphical representation of a probabilistic model of hospital patient fatality rates after accidental falls in children. A child may sustain a short fall with probability *f*_*s*_, or a long fall with probability *f*_*l*_. The child may develop eventually fatal injuries with probability *d*_*s*_ after a short fall, and *d*_*l*_ after a long fall. After a fall, all children are brought to the hospital, except after short nonfatal falls, in which case this occurs with probability *h*.

In addition to direct calculations, we conducted numerical simulations with the Python programming language, using the open-source scientific libraries NumPy, matplotlib, seaborn, and Jupyter. The open-source code is freely available on the GitHub website^1^.

## 3. Results

Using mathematical modeling and numerical simulations, we investigate two factors that may influence the hospital patient fatality rate after accidental falls: the consultation rate at the hospital after short household falls and the age of the child. Conclusions about the truthfulness of stories of accidental falls should take these factors into account.

### 3.1. Influence of Hospital Consultation Rate on Hospital Patient Fatality Rates

We perform both analytical and numerical computations.

#### 3.1.1. Analytical results

We can directly calculate the expected number of patients in the hospital in each category: short nonfatal, short fatal, long nonfatal, long fatal. The key parameter of the model, *h*, only appears in the first category. The four expressions are shown in Table 1 (*N* is the total size of the population).

**Table 1:** Expected number of patients in each category.

|  | short falls | long falls |
| --- | --- | --- |
| nonfatal | $N f_s(1 - d_s)h$ | $N f_l(1 - d_l)$ |
| fatal | $N f_s d_s$ | $N f_l d_l$ |

From these expressions, we derive the *hospital patient fatality rates* after short falls (*r*_*s*_) and long falls (*r*_*l*_):

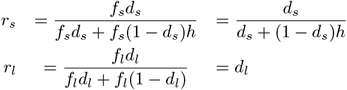

In this model, the *hospital patient fatality rate* after long falls, *r*_*l*_, equals the *actual fatality rate* after long falls, *d*_*l*_. This is due to the fact that all long falls involve a consultation at the hospital.

On the other hand, the hospital patient fatality rate after short falls, *r*_*s*_, depends on the key parameter *h*, the probability that a short fall will lead to a consultation at the hospital. It is easy to prove that *r*_*s*_(*h*) is a decreasing function of *h*, with *r*_*s*_(0) = 1 and *r*_*s*_(1) = *d*_*s*_. If *h* = 1, all short falls are seen in the hospital and *r*_*s*_ = *d*_*s*_. At the other extreme, if *h* = 0, nonfatal short falls are never seen in the hospital, therefore, the hospital patient fatality rate after short falls would be 1. Between these two extremes, when *h ∈* (0, 1), the hospital patient fatality rate after short falls *r*_*s*_ may take any value in the range (*d*_*s*_, 1), as shown in Figure 2.

**Figure 2:**
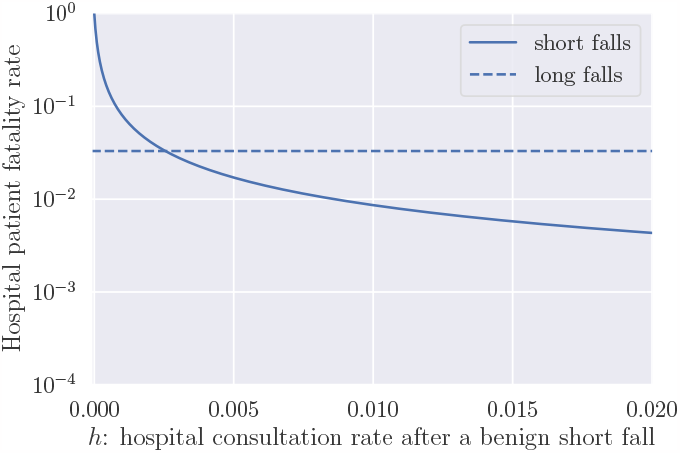
Hospital patient fatality rate after short falls (*r*_*s*_), as a function of the probability that a child sustaining a benign short fall will be brought to the hospital (*h*). A logarithmic scale is used for the *y* axis. The fatality rate after long falls, which does not depend on the *x* axis variable, is shown as a dashed line. Realistic parameters were used for this figure (see later in this article): *f*_*s*_ = 0.5, *f*_*l*_ = 9.3 *×* 10^−5^, *d*_*s*_ = 8.7 *×* 10^−5^, *d*_*l*_ = 3.3 *×* 10^−2^.

We observe that the hospital patient fatality rates after accidental falls may differ significantly from the actual fatality rates, depending on the height of the falls. The rates depend critically on the denominator, especially because the vast majority of benign short falls do not lead to a consultation at the hospital:

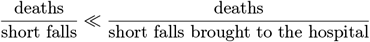

For small values of *h*, it then becomes possible for the hospital patient fatality rate to be *higher* after short falls than long falls, which represents an apparent paradox:

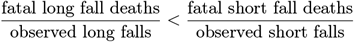

The *actual* fatality rate is, of course, significantly higher after long falls versus short falls.

The hospital patient fatality rate is extremely sensitive to *h*, which is, of course, difficult to estimate reliably. It depends on many factors: the region, socioeconomic status, cultural factors, the state of public healthcare, the period of the year, and others. We can expect this parameter to be very small, of the order of 1% or 0.1%. An estimation of *h* based on real data is proposed later in this paper.

A graphical representation of this apparent paradox is shown in Figure 3.

**Figure 3:**
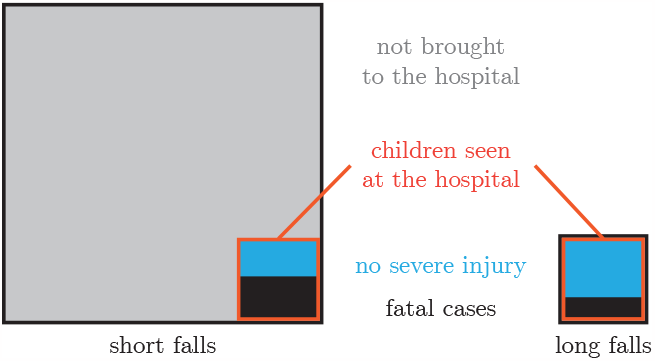
Graphical representation of a statistical paradox explaining why the hospital patient fatality rate may be higher after short falls versus long falls. *Left*: the large gray square represents all short falls within a given population of children. The red-bordered square represents the children brought to the hospital after the falls. The blue rectangle represents the non-fatal cases, whereas the black square represents the fatal cases. We assume that all eventually fatal cases lead to a consultation at the hospital. *Right*: the red-bordered square represents all long falls within the same population. They are much rarer than short falls. However, here, all long falls lead to a consultation at the hospital. The black area relative to the encompassing black-bordered square is higher with long falls versus short falls, but this trend is reversed when comparing to the red squares, which represent the populations observed in hospitals. The shapes are not to scale; they merely illustrate a mathematical pattern.

#### 3.1.2. Numerical computations: parameter estimation

We also conducted numerical simulations of our model to estimate the distribution of the hospital patient fatality rate after short falls and long falls.

Following [10], we used realistic parameters to approximately match Chadwick’s data.

The Trauma Center at Children’s Hospital San Diego served a population of about 2.2 million people around 1985 (the San Diego County) [11]. About 4% of the population was aged 0–4, and 15% was aged 0–15 [12]. About 1% of the population was less than 12 months. The number of children less than 4 years old in the area was therefore about *N* = 88, 000.

The study’s duration was 44 months, or *T* = 3.67 years.

Chadwick’s article omits key information: the age of the child in each fall group (short falls, medium falls, long falls), although the total number of child in each age group is given. Following [10], we can make reasonable estimates based on a few plausible assumptions, for example that most infants fell from low heights, whereas children falling from medium heights (playground) were on average somewhat older. We find that, whereas 118 long falls were observed in Chadwick’s study in the 0–15 age group, there may have been about 30 long falls in the 0–4 age group. We also assume this number represents virtually all long falls of children in the population served by the hospital during the time period (except the cases where the child died at the scene, which would further increase the long fall fatality rate).

Therefore, we estimate the long fall rate among children 0–4 to be *f*_*l*_ = 30*/*(88, 000 *× T* ) *≃* 9.3 *×* 10^−5^.

Similarly, we estimate the observed number of short falls at the hospital in the 0–4 age group to be 100.

To estimate the hospital consultation rate, we use data from [6] that showed that about half of all infants sustained a short fall during their first year of life, so we take *f*_*s*_ = 0.5.

Therefore, *f*_*s*_ × *h* × *N* × *T* = 100, so *h* = 1.2×10^−3^.

Finally, we estimate the death rates. For long falls, there was one death among roughly 30 long falls in the 0–4 age group, so *d*_*l*_ = 1*/*30 *≃* 3.3 *×* 10^−2^. For short falls, there were 7 deaths, so *d*_*s*_ = 7*/*(*N × f*_*s*_ *× T* ) *≃* 1*/*12000 *≃* 8.7 *×* 10^−5^.

#### 3.1.3. Numerical results

With this set of parameters matching Chadwick’s own data, we ran 1000 probabilistic simulations of the model, and we computed the hospital patient fatality rate for short falls and long falls (see Figure 4). We found that the hospital patient fatality rate was higher after short falls than long falls in 72% of our simulations (in Figure 4B, this represents the relative area under the curve to the right of the vertical black line). Therefore, this observation in Chadwick’s data is not illogical, as it can be perfectly explained by our simple model.

**Figure 4:**
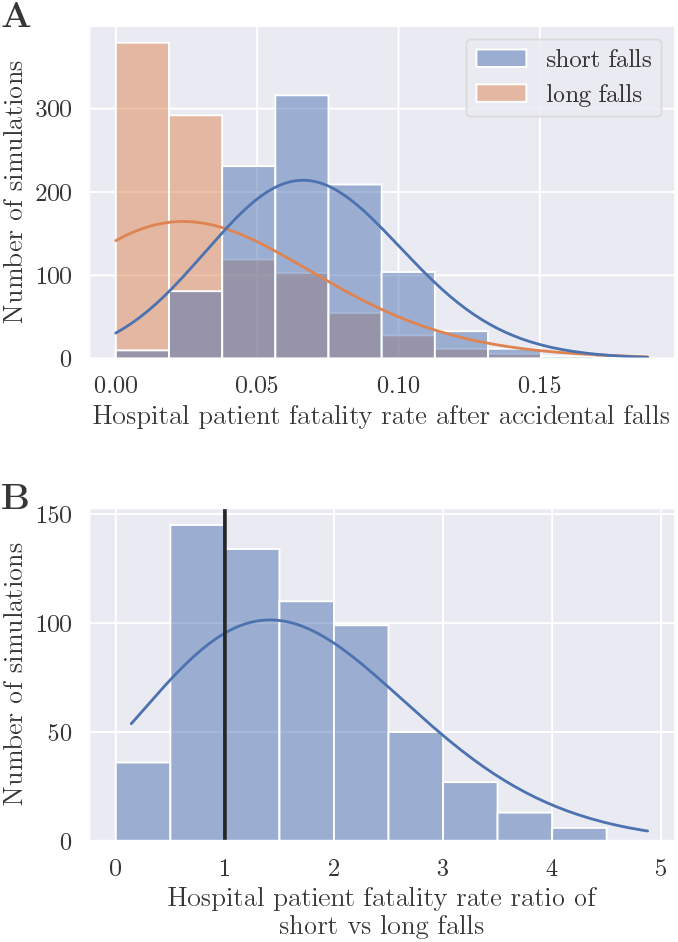
Numerical simulations of hospital patient fatality rates after short falls and long falls. **A**. Histogram of hospital patient fatality rates after short falls (blue) and long falls (orange), estimated with 1,000 numerical simulations of populations of 88,000 children. **B**. Histogram of the ratio between the hospital patient fatality rate after short versus long falls, using the same simulations as in panel A. The vertical black line shows the value 1, i.e., equal hospital patient fatality rate after short falls and long falls. The following values were used for this figure: *f*_*s*_ = 0.5, *f*_*l*_ = 9.3 *×* 10^−5^, *d*_*s*_ = 8.7 *×* 10^−5^, *d*_*l*_ = 3.3 *×* 10^−2^, *h* = 1.2 *×* 10^−3^.

### 3.2. Influence of the Age of the Child

The consultation rate after benign short falls suffices to explain the apparently illogical observation in [8] that the hospital patient fatality rate was higher after short falls than long falls. However, other confounding variables may also lead to this observation. One of them is the age of the child, which influences both the relative frequencies of short vs long falls and the hospital patient fatality rate after falls.

Premobile infants are much more likely to fall from low heights (falling from the arms, the changing table, the sofa, the bed), whereas long falls are more common in older children than infants. On the other hand, the fatality rate after falls is likely to be higher in infants than older children, for a given height, since younger children are more fragile than older ones [13]. These considerations prove that other factors can also contribute to producing the seamingly paradoxical, but actually logical result observed in [8].

## 4. Discussion

In [8], Chadwick drew a strong general conclusion from his data: *“when children incur fatal injuries in falls of less than 4 feet [1*.*20 meters], the history is incorrect*.*”* This study has been greatly influential in the child abuse pediatrics community, being cited more than 300 times in the past 30 years. The guidelines of the French national health authority on the diagnosis of shaken baby syndrome cite this study and others to reject all histories of short falls in infants presenting with intracranial and intraocular hemorrhage [14]. In a videotaped conference^2^, the president of the working group of these guidelines said: *“this article showed that the only children dying after falls were those falling from less than one meter, so the only explanation was obviously violence*.*”*

As we have shown in this article, this conclusion cannot be drawn from the data because at least one confounding variable (the proportion of short benign falls being brought to the hospital) explains the discrepancy between the hospital patient fatality rate after short versus long falls.

Chadwick’s article mentions associated injuries in some of the fatal short fall cases: 2 cases with old fractures, 2 cases with bruises on the trunk or extremities, 2 cases with genital injury, 2 cases with two head impact sites. There were two cases with no associated injury. Because of the existence of these associated injuries, it is plausible that at least some of these fatal cases involved violence. The discrepancy between the reported clinical history and the severity of injuries has long been interpreted as a marker of child abuse. Perpetrators of child abuse, when bringing an injured child to the hospital, may report a false history of short falls to avoid confessing a crime and being liable for criminal prosecution.

Nevertheless, even excluding these five cases with associated injuries, the number of fatal short falls (two) remains higher than after long falls (one). This discrepancy is still explained by the confounding variable, the proportion of short benign falls being brought to the hospital. In other words, our model by no means purports to prove that *all* histories of fatal short falls are genuine, but that *some* may be, even if the hospital patient fatality rate is higher than for long falls.

Other factors were not appropriately taken into account by [8]. For example, the study did not provide sufficient data regarding the age of the children. Most long falls are probably experienced by older children who can climb.

Note that some children falling from heights may be found dead on the scene and never be brought to the hospital, so that the hospital patient fatality rate may be even lower than the true long fall fatality rate.

## 5. Conclusion

We have shown that the observation of a hospital patient fatality rate unexpectedly higher after short falls versus long falls cannot be used to draw general conclusions about the truthfulness of histories of accidental falls. Although suspicions of nonaccidental trauma can be warranted, for example in the presence of unrelated or earlier injuries, paradoxical results should not systematically be interpreted as proofs of abuse, especially when statistical issues are at play. Our study showed that even basic statistical models can explain unexpected statistical results in epidemiological studies such as those concerning accidental falls.

### CRediT authorship contribution statement

**Cyrille Rossant:** Conceptualization of this study, Methodology, Software, Writing. **Leila Schneps:** Conceptualization of this study, Methodology, Writing.

## Data Availability

All data produced in the present study are available upon request to the authors

## Footnotes

1 https://github.com/rossant/paper-chadwick

2 https://www.youtube.com/watch?v=KjK6j2SoxXw

